# Gradient-guided adapter merging for neuroimaging vision-language models

**DOI:** 10.64898/2026.07.18.26358397

**Authors:** Subhrangshu Bit, Osman Berke Guney, Shuyue Jia, Vijaya B. Kolachalama

**Author notes:** Corresponding author: V.B.K.

## Abstract

Automated interpretation of neuroimaging studies requires simultaneous assessment of multiple imaging evidence variables, each tied to distinct anatomical structures. Vision-language models (VLMs) offer a unified framework for multi-task analysis, but adapting pre-trained VLMs remains challenging. Full fine-tuning is computationally prohibitive, and joint multi-task training requires simultaneous access to all task data, which is often infeasible in clinical settings. Although model merging enables multi-task composition without joint re-training, existing methods focus on post-hoc algorithms with limited extension to VLMs and minimal application to neuroimaging. Here, we present GRadient-guided Adapter Merging (GRAM), a layer-selective low-rank adaptation (LoRA)-based fine-tuning and merging framework for multi-task neuroimaging visual question-answering (VQA). GRAM uses a gradient ratio that contrasts class-specific gradients to identify task-discriminative layers, and applies subspace-constrained projected gradient descent to restrict LoRA updates to directions consistent with the geometry of the pre-trained model. We leveraged a structured VQA benchmark, developed from the National Alzheimer’s Coordinating Center (NACC) dataset, that pairs multisequence brain MRI studies with question-answer pairs across clinically relevant imaging evidence variables. Experiments on the VQA benchmark showed that GRAM outperformed or matched all-layer LoRA fine-tuning and a standard merging baseline while reducing inter-task interference during merging, and approached or surpassed the performance of joint multi-task training without joint re-training.

## 1 Introduction

Vision-language models (VLMs) trained on largescale paired image-text corpora have become integral to a range of tasks in modern image analysis [1, 2]. These models exhibit remarkable transferability across diverse tasks, including medical imaging, through fine-tuning. However, such models often require modality-specific adaptation to address the inherent differences among medical imaging modalities, such as the various types of MRI sequences. Due to the limited scale of medical data, most studies have resorted to adapter-based fine-tuning as a parameter-efficient strategy, allowing each specialist module to acquire task-specific representational capabilities while retaining the shared knowledge of the pre-trained backbone. Nevertheless, developing a unified model that generalizes across multiple imaging modalities and tasks remains challenging. Training a single medical vision-language model typically demands large, heterogeneous datasets [3, 4], which are often impractical to curate in clinical contexts. An attractive alternative is model merging, i.e., combining independently fine-tuned models into a single network without additional training. Most existing merging methods employ post-training fusion rules that disregard the adaptation stage of individual tasks. This static nature limits their ability to capture fine-grained task characteristics, leading to parameter interference and suboptimal generalization.

The widespread post-training deployment of large VLMs across diverse clinical applications has led to an ecosystem of independently fine-tuned specialist models, each trained on proprietary datasets held by different institutions. This fragmentation motivated a modular approach based on model merging: rather than centralizing data, individual institutions fine-tune their own adapter on local data and contribute only the resulting model weights. This enables collaboration without data sharing, which is particularly important in healthcare, where regulatory constraints, patient consent requirements, and institutional data governance policies make sharing raw imaging data across sites difficult or infeasible. Model merging therefore offers a privacy-preserving mechanism for multi-task generalization, as the merged model inherits complementary clinical knowledge from each contributor without requiring patient-level data to leave the originating institution. Several paradigms exist for this kind of privacy-preserving collaboration. Parameter averaging [5] and task arithmetic [6], which encoded fine-tuning as a weight-space offset and added task vectors to form a multi-task model, have established the foundational composition paradigms. Despite a growing body of algorithmic refinements, these methods treated the fine-tuning stage as fixed and applied merging posthoc. A key insight is that the geometric structure of the fine-tuned task vectors is one of the primary determinants of merging success [7], suggesting that adapter training can be as important as the merging procedure itself.

In neuroimaging, these challenges are further compounded. Brain MRI studies are multi-sequence volumetric acquisitions that differ fundamentally in spatial structure and contrast from 2D natural images on which VLMs are generally pre-trained. Different imaging findings, such as hemorrhage, atrophy and white matter lesions, engage spatially distinct anatomical regions, suggesting that their discriminative signal is unlikely to be uniformly distributed across transformer layers. Yet most existing adapter fine-tuning approaches apply low-rank adapters (LoRA) [8] uniformly across attention layers, ignoring this heterogeneity, and, to our knowledge, model merging has not been systematically studied for neuroimaging VLMs.

To address these limitations, we proposed GRadient-guided Adapter Merging (GRAM), a data-adaptive fine-tuning and merging framework that (i) identifies which layers carry task-discriminative signal through a class-contrastive gradient criterion, (ii) restricts LoRA fine-tuning to those layers using subspace-constrained gradient descent, and (iii) composes the resulting task vectors into a unified multitask VLM without any joint re-training or data sharing.

### 1.1 Related work

#### 1.1.1 Sparse adaptation of large pre-trained models

The prohibitive cost of full fine-tuning of large pre-trained models has motivated a rich line of parameter-efficient fine-tuning (PEFT) methods that update only a small subset of parameters while keeping the pre-trained backbone frozen. These include adapter layers [9], prefix tuning [10], prompt tuning [11], and low rank updates [8]. LoRA in particular has become the dominant PEFT approach for large-scale models due to its inference efficiency and compatibility with model merging pipelines [12, 13]. A complementary direction is sparse fine-tuning, which updates a small subset of parameters rather than a low-rank subspace of all parameters. [14] allowed adaptive allocation of parameters among layers that are important for the task. Sparse fine-tuning strategies exploit gradient-based importance criteria to identify which parameters to update [15, 16], often leveraging the Fisher information matrix (FIM) to score parameter sensitivity [17, 18]. TaLoS updated only the least sensitive parameters, demonstrating that this promoted function localization and reduced task interference during merging without requiring explicit model linearization [19]. Attention-only finetuning similarly restricts updates to attention layers and shows that this implicitly induces kernel-like behavior conducive to weight disentanglement [20]. Sparse masking techniques further exploit parameter subnetworks for continual and multi-task learning [21, 22]. Our work differs from these methods in that we do not select parameters based on absolute sensitivity scores or structural constraints; instead, our method employs a class-contrastive criterion that identifies which layers are disproportionately activated by radiological as opposed to normal signals.

#### 1.1.2 Model merging

The goal of model merging is to unify independently fine-tuned models into a single multi-task model without joint re-training. Early approaches focus on simple parameter averaging [5, 23], and exploit mode connectivity between solutions in the loss landscape to improve compatibility before averaging [24]. Alignment-based methods reduce interference by matching neurons or features across models prior to merging [25, 26]. Re-weighting schemes exploit the Fisher information matrix [27], or data-driven inner product matrices [28], but incur high memory costs. Task arithmetic established a dominant paradigm: a task vector is defined as the parameter offset induced by fine-tuning, and multiple task vectors are summed with scalar coefficients to form a multi-task model [6]. TIES-merging [29] refined this by pruning low-magnitude task vector entries and resolving sign conflicts before summation. SVD-based subspace selection identifies task-relevant directions in weight space [30]. More recent work introduces dynamic merging through data-adaptive routing [31] and hypernetworks to generate instance-specific weights on the fly [32] at the cost of requiring a separately pretrained generator. An emerging body of work redirects attention from post-hoc fusion to the fine-tuning stage itself. The effectiveness of task arithmetic has been shown to depend critically on weight disentanglement, the property that a task vector modifies behavior only within its own data support [7], and linearized fine-tuning has been shown to amplify this property. Linearized LoRA [33] fine-tunes adapters in the tangent space of the pre-trained model at a fraction of the computational cost of full linearization.

#### 1.1.3 VLMs for neuroimaging

Large VLMs have demonstrated strong generalization in medical image analysis [2, 1], including domain specific VLMs for radiology report generation for chest X-rays [34], MRI [4, 3], and pathology [35]. In neuroimaging specifically, deep learning methods have been applied to tumor segmentation [36], dementia classification [37]. However, adapting large VLMs to the specific structure of multi-sequence brain MRI remains largely unexplored. Existing medical VLM adaptation works focus predominantly on 2D single-modality imaging and do not address the challenge of multi-sequence volumetric MRIs. Critically, to our knowledge, no prior work has applied model merging to VLMs for neuroimaging, leaving the multi-task composition problem unaddressed in this domain.

### 1.2 Contributions

The main contributions of this paper are summarized below:

- We proposed and validated a class-contrastive layer importance metric, the gradient ratio, for selective fine-tuning of VLMs on neuroimaging tasks.
- We introduced a geometry-aware fine-tuning strategy that applies subspace-constrained gradient descent to the LoRA factors, restricting task-specific updates to the column space of the pre-trained weights. Keeping the task vectors aligned with the geometry already encoded in the backbone yields merge-friendly adapter training.
- We established, to our knowledge, the first systematic evaluation of model merging for VLMs operating on multi-sequence brain MRI, and demonstrated that GRAM narrows the gap to the joint multi-task training ceiling without any data sharing or joint re-training.

## 2 Materials and methods

### 2.1 Study population

We obtained access to a large-scale neuroimaging and demographic dataset from National Alzheimer’s Coordinating Center (NACC) (Table 1). As in [4], we followed the construction of a visual questionanswering (VQA) benchmark from cases in the NACC cohort with documented imaging evidence. Each case includes multiple MRI sequences paired with the patient’s context and categorical QA annotations derived from structured imaging variables. Specifically, we combined NACC variables into four broader sets of tasks namely: Hemorrhage (IMAGMICH and IMAGMACH), temporal atrophy (HIPPATR), frontal atrophy (MRFTLD), and white matter hyperintensity (INFWMH, IMAGEWMH). For patient context, we formed a single sentence as a prompt, including age, sex and years of education. For each task, six question types were defined to improve linguistic robustness.

**Table 1:**
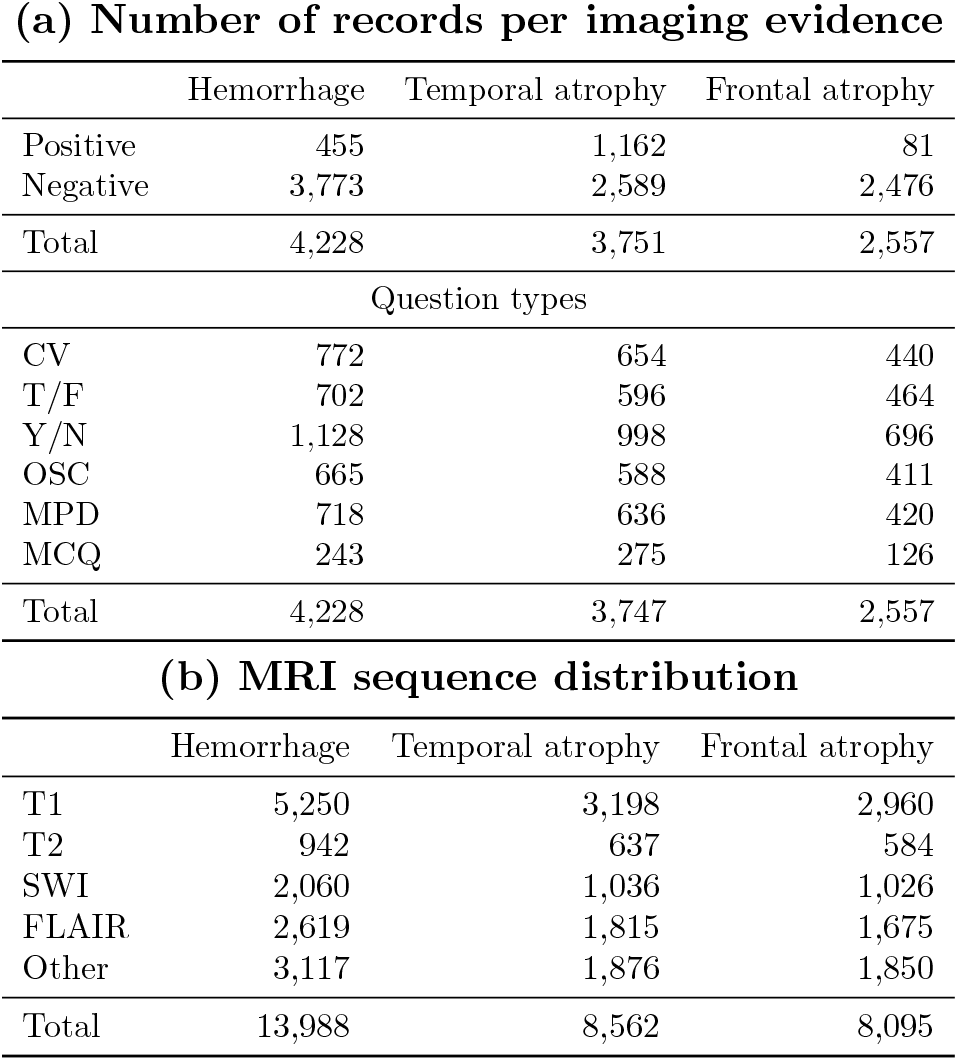
Study population. Visual question-answering dataset constructed from NACC. Six different question types were used — CV: Choice Verification; T/F: True/False; Y/N: Yes/No QA; OSC: Opposite Statement (Contradiction); MPD: Minimal Pair Discrimination; MCQ: Multiple Choice QA.

|  | Hemorrhage | Temporal atrophy | Frontal atrophy |
| --- | --- | --- | --- |
| Positive | 455 | 1,162 | 81 |
| Negative | 3,773 | 2,589 | 2,476 |
| Total | 4,228 | 3,751 | 2,557 |
| Question types |  |  |  |
| CV | 772 | 654 | 440 |
| T/F | 702 | 596 | 464 |
| Y/N | 1,128 | 998 | 696 |
| OSC | 665 | 588 | 411 |
| MPD | 718 | 636 | 420 |
| MCQ | 243 | 275 | 126 |
| Total | 4,228 | 3,747 | 2,557 |

(b) MRI sequence distribution
|  | Hemorrhage | Temporal atrophy | Frontal atrophy |
| --- | --- | --- | --- |
| T1 | 5,250 | 3,198 | 2,960 |
| T2 | 942 | 637 | 584 |
| SWI | 2,060 | 1,036 | 1,026 |
| FLAIR | 2,619 | 1,815 | 1,675 |
| Other | 3,117 | 1,876 | 1,850 |
| Total | 13,988 | 8,562 | 8,095 |

### 2.2 Methodology

Let 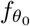 denote a pre-trained VLM with *L* layers, parameterized by 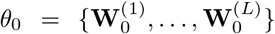, where 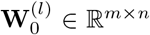 represents the weight matrix of the *l*-th layer. Let *T* = {*T*_1_, …, *T*_*N*_} denote a collection of *N* neuroimaging tasks, where each task corresponds to a clinically relevant imaging finding, such as hemorrhage or atrophy. Each task *T*_*t*_ is associated with a dataset 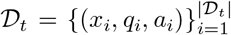, where *x*_*i*_ denotes a multi-sequence brain MRI study represented as a set of video inputs, and (*q*_*i*_, *a*_*i*_) denotes the corresponding question-answer pair. The goal is to obtain a single unified model 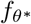 that performs competitively across all tasks in *T*, without requiring joint multitask training.

#### 2.2.1 Background

This subsection reviews parameter-efficient finetuning and model merging concepts, which provide the foundation for integrating task-specific adaptations into a unified VLM without joint multi-task training.

##### Parameter-efficient fine-tuning

Full finetuning of a pre-trained VLM separately for each task is computationally prohibitive and can lead to catastrophic forgetting of pre-trained visual-language representations [8, 38]. LoRA mitigates this issue by freezing the pre-trained weights and learning task-specific updates in a low-rank subspace [8]. For task

*T*_*t*_, given a weight matrix 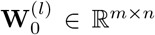 at layer *l*, LoRA parameterizes the task-adapted weight as

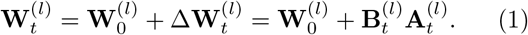

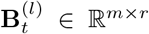 and 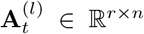 are trainable lowrank matrices for task *T*_*t*_, with rank *r* ≪ min(*m, n*). The pre-trained weight 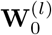 remains frozen during training, and only the low-rank factors 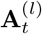 and 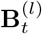 are updated. Applying LoRA to a subset of layers *M* ⊆ 1, …, *L* therefore produces task-specific lowrank updates 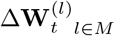, while the weights of the non-adapted layers remain unchanged. The resulting task-adapted model parameters can be written as

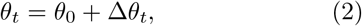

where Δ*θ*_*t*_ collects all LoRA-induced updates for task *T*_*t*_ across the adapted layers, with 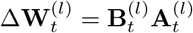 for *l* ∈ *M* and no update applied to layers outside *M*.

##### Model merging

Task arithmetic [6] frames the composition of multiple independently fine-tuned models into a single unified model, without additional training, as arithmetic over task vectors. For task *T*_*t*_, the task vector is defined as *τ*_*t*_ = *θ*_*t*_ ™ *θ*_0_, i.e., the difference between the task-adapted parameters and the original pre-trained parameters.^1^ A multi-task model is then constructed by adding a weighted combination of task vectors to the pre-trained model,

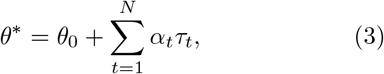

in which *α*_*t*_ is a scalar coefficient controlling the contribution of task *T*_*t*_ to the merged model. A key challenge in this formulation is that task vectors learned independently may overlap or conflict in weight space. Direct additive composition can therefore introduce destructive interference, which may degrade task-specific performance relative to the corresponding individually fine-tuned models. This motivates fine-tuning strategies that encourage task-specific updates to be more amenable to merging.

#### 2.2.2 GRAM

We introduce GRAM to improve the merge-compatibility of task-specific LoRA updates by controlling both their layer-wise support and their optimization geometry. GRAM first identifies, for each task, the layers whose gradients are most selectively driven by task-discriminative radiological signal rather than domain-generic MRI structure. It then constrains the LoRA updates within the selected layers through projected gradient descent, encouraging the resulting task vectors to remain aligned with the geometry of the pre-trained model while reducing destructive interference during merging. An overview of GRAM is shown in Fig. 1, and the details of its two main components are described below

**Figure 1:**
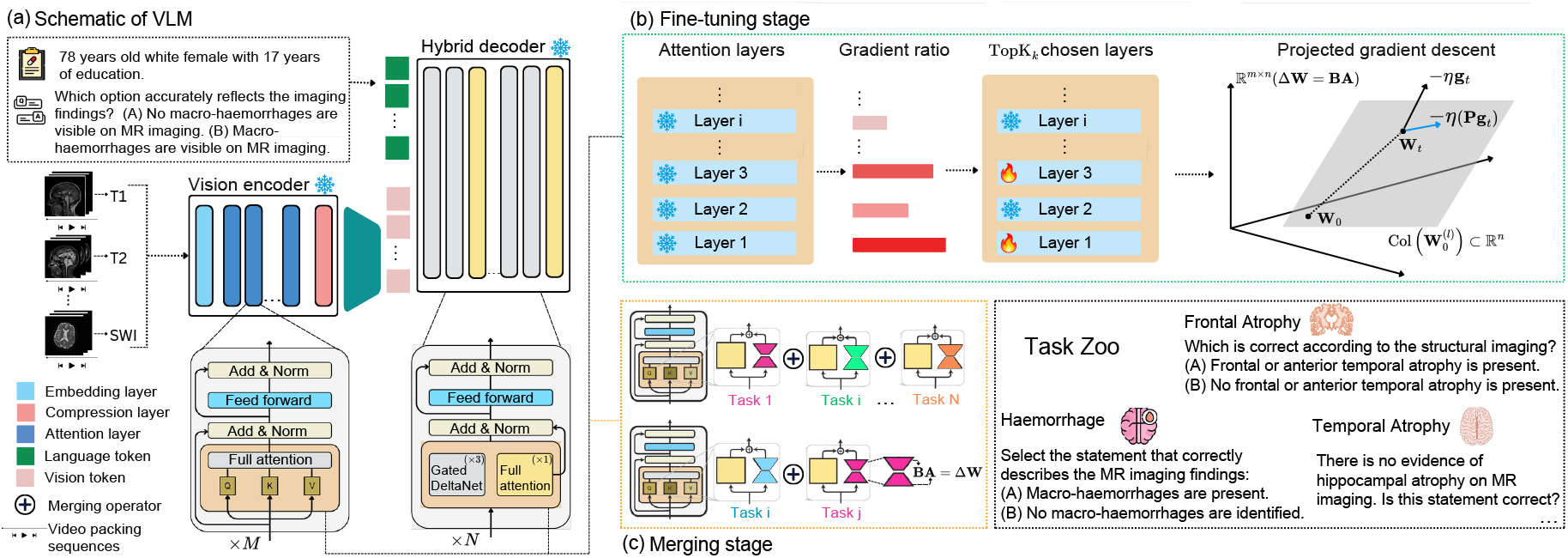
Overview of GRAM. (a) Each study containing multi-sequence brain MRI was formatted as video representation, which were given as input to a VLM with patient demographics and a prompt. (b) For each task, a calibration pass computed class-specific gradient magnitudes per layer. The gradient ratio identified the TopK_*k*_ layers discriminative of each task’s radiological signal. These layers were fine-tuned using projected gradient descent. (c) LoRA adapters were applied exclusively to the selected layers, yielding sparse, task-selective experts. These task-specific adapters are merged into a single unified model without additional training.

##### Layer importance signal

A key design choice in LoRA fine-tuning is where to apply the trainable low-rank updates. Standard LoRA configurations commonly apply adapters uniformly across transformer layers [8]. However, recent studies suggest that this strategy can be suboptimal, as different layers and modules may contribute unequally to downstream adaptation [14, 39]. We revisited this design choice in the neuroimaging setting and hypothesized that gradient information accumulated during an initial calibration probe could identify which layers are most sensitive to each task. For task *T*_*t*_, the pre-trained parameters *θ*_0_ are treated as differentiable during the calibration probe, but no parameter update is applied. Given a calibration mini-batch ℬ ⊂ *D* _*t*_, the gradient of the task loss with respect to the weight matrix **W**^(*l*)^ at layer *l* is computed As 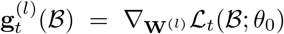. The Frobenius norm 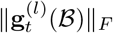 quantifies the magnitude of the parameter change induced at layer *l* by the task-specific training signal, with larger values indicating layers whose representations are more strongly affected by the task. However, in neuroimaging VLM adaptation, large gradient magnitudes may primarily reflect domain adaptation to the shared visual structure of brain MRI rather than sensitivity to the radiological finding of interest. This issue is further amplified by class imbalance, since normal cases are often substantially more frequent than cases with positive imaging findings. To distinguish task-discriminative radiological signal from domain-generic MRI adaptation, the calibration data for each task are separated into positive and negative subsets. Let 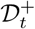 and 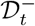 denote the subsets of *D* _*t*_ corresponding to the radiological signal class and the normal class, respectively. The class-specific gradient magnitudes at layer *l* are defined as

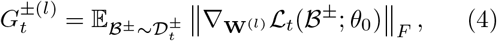

where + and correspond to the radiological signal and normal classes, respectively. In absolute terms, these quantities can remain difficult to compare across layers, since both positive and negative examples may induce large gradients in layers responsible for general MRI representation learning. Therefore, layer importance is estimated from the relative contrast between these quantities, as defined by the gradient ratio (GR):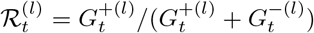. The score 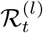 quantifies the relative contribution of the radiological signal class to the total class-specific gradient magnitude at layer *l*. 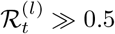 indicates that positive examples induce larger loss-driven parameter changes than normal examples, suggesting that layer *l* is selectively sensitive to the task-relevant radiological signal. In contrast, 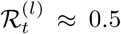 indicates comparable gradient responses for positive and normal examples, suggesting that the layer primarily captures domain-generic MRI structure rather than task-discriminative representations. Given the gradientratio scores 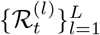 for task *T*_*t*_, the adapted layer set is defined by selecting the *k* layers with the largest scores:

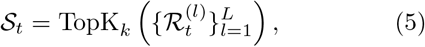

where TopK_*k*_ (·) returns the indices of the *k* highest-scoring layers, and *k* is a hyperparameter controlling the number of adapted layers. We additionally constructed class-specific layer subsets: the positive class is assigned the *k* layers with the highest 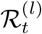 scores, while the negative class is assigned the *k* layers with the lowest scores, corresponding to the max- and min-TopK of the positive-class gradient ratio for a given task. LoRA adapters are applied only to layers in *S*_*t*_, while all other layers remain unchanged. The layer-wise transformation can therefore be written as

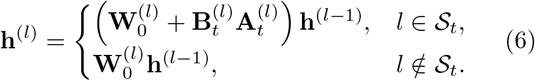

This produces a sparse, task-selective LoRA adapter in which the update for task *T*_*t*_ is restricted to the layers most strongly associated with its task-discriminative radiological signal.

##### Projected gradient descent

While GR identifies which layers should receive LoRA updates for each task *T*_*t*_, merge performance also depends on how the updates within those layers are optimized. Stochastic gradient descent on the LoRA matrices 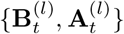 optimizes task-specific updates without an explicit geometric constraint within the parameter space of each selected layer, so the resulting task vector *τ*_*t*_ may occupy directions that interfere with task vectors learned for other tasks. Prior work has often used task-vector orthogonality as a proxy for merge compatibility of these task vectors [6, 29]. The idea is that near-orthogonality between task vectors reflects greater disentanglement in weight space, thereby reducing the likelihood of destructive interference during merging. However, this criterion is most directly justified under linearized approximations of the network [19], and may be less reliable for non-linear architectures, where merge compatibility between models can be better reflected by alignment in layer activations [30]. To enhance the compatibility of task vectors for merging, we constrain the geometry of task-specific LoRA updates during finetuning, guiding them toward transformations that remain aligned. Specifically, for each task *T*_*t*_, the constrained fine-tuning objective is:

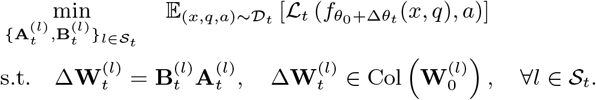

The intuition behind this constraint is that the frozen pre-trained weights define a shared geometric reference frame across all task-specific adaptations. For each selected layer *l*, the base weight matrix 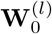 defines the transformations already supported by the pre-trained model, with its column space representing the output directions actively used by that layer. Since every task is adapted from the same pre-trained model, constraining each task-specific LoRA update to remain aligned with this shared subspace encourages the resulting task vectors to be expressed relative to a common layer-wise geometry. This may reduce arbitrary task-specific deviations in weight space and encourages task vectors that are more compatible when combined during merging. This subspace constraint is implemented through projected gradient descent. For each selected layer *l* ∈ *S*_*t*_, the SVD of the frozen pre-trained weight matrix is computed as:

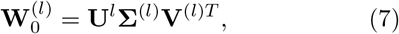

where **U**^(*l*)^ ∈ ℝ^*m×m*^ and **V**^(*l*)^ ∈ ℝ^*n×n*^ are orthogonal matrices containing the left and right singular vectors, respectively, and **Σ**^(*l*)^ contains the singular values 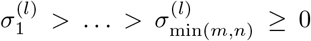 along its diagonal. The decomposition is used to define the following projection operator corresponding to the *e*-largest singular values:

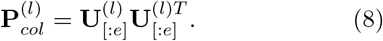

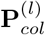 projects onto the column space of 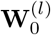. At each training step, the gradient of the task loss with respect to the LoRA update 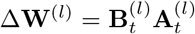 is projected before descent.

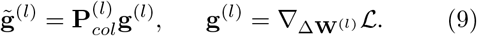

The projected gradient is back-propagated through the LoRA factorization via:

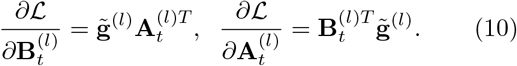

requiring no modification to the LoRA architecture or any post-trained merging algorithm.

## 3 Experiments

### 3.1 Dataset

All experiments were conducted on NACC-VQA benchmark. Each study comprised multiple MRI sequences, encoded as a corresponding set of interleaved synchronized video inputs to the vision tower of Qwen3.5-4B. Question-answer pairs were formulated across four imaging-evidence tasks: Hemorrhage, Temporal atrophy, Frontal atrophy, and White matter hyperintensity, with question-formats including multiple-choice, yes/no, true/false, choice verification, minimal pair discrimination, and opposite statement. The dataset was split into training, validation, and held-out test sets stratified by different question types and imaging evidence while ensuring there was no patient-level overlap. To prepare multi-sequence MRI inputs for VLM, each 3D volume was converted to a fixed length sequence of 2D frames through video packing. All volumes were loaded as grayscale arrays and resampled in-plane to a uniform spatial resolution of *H* = *W* = 128 pixels using linear interpolation. Per-volume intensities were normalized using min-max scaling. To retain the highest resolution, the primary slice direction was inferred from the DICOM header voxel spacing: the axis with the largest voxel spacing was identified as depth and the volume array was permuted accordingly. Up to 32 frames were then sampled uniformly along the depth dimension across the full extent of the volume. All-zero slices were removed before sampling. For VLM input, all sampled frames per volume per modality were concatenated into a single contiguous video-style sequence. When multiple volumes existed for the same modality within a visit, each video is separated using a tag of the modality name and the volume count. The language tower of the VLM was contextualized using the patient’s age, sex and education, followed by the question.

### 3.2 Implementation details

#### 3.2.1 Model

All experiments used Qwen3.5-4B as the base VLM. The vision tower encoded the interleaved multisequence video frames, while the language block combined these visual embeddings with patient context to answer VQA prompts. The base model weights remained frozen throughout all experiments; only LoRA adapter parameters were updated during finetuning.

#### 3.2.2 LoRA configuration

LoRA adapters were applied to the attention projection matrices (**W**_*q*_, **W**_*k*_, **W**_*v*_, **W**_*o*_) of the transformer layers selected under each configuration. All adapters used rank 8, scaling factor 16, and dropout 0.10. We used AdamW with warmup=0.05 and a cosine learning rate. Learning rates and weight decay were task-specific due to differences in class frequencies. Each task was fine-tuned for 3 epochs with early stopping and gradient accumulation.

#### 3.2.3 Gradient ratio

The gradient ratio was computed over a single probe epoch prior to main fine-tuning stage. Class-balanced mini-batches of size 25 were drawn separately from the imaging evidence 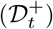 and normal 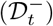 subsets of each task’s validation split. Gradient Frobenius norms were accumulated over the LoRA adapter factors at each layer and averaged across batches to estimate 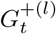 and 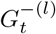. We then select the top-K layers for both + and ™ classes of each task.

#### 3.2.4 Projected gradient descent

The subspace constrained gradient descent was applied during the first 100 fine-tuning steps, after which training continued with standard gradient descent. This warm start anchored the early adapter trajectory within the structured subspace of 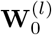, while the subsequent unconstrained phase allowed the adapter to recover task-discriminative signals that may lie outside the projected space. In addition, instead of random Gaussian initialization for **A**^(*l*)^, the LoRA low-rank factors were initialized using top-90% singular vectors of 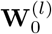.

### 3.2.5 Merging

For linear merging, the scaling coefficient *α* was set to 1.0 for all tasks. For TIES merging, we used the recommended hyperparameters setting pruning threshold at 0.2 and the scaling values *α*=1.0. All merging was performed in a single forward pass with no additional training.

### 3.3 Computing infrastructure

All experiments were performed on an NVIDIA L40S graphics card with 48GB memory. Training each task required roughly 3 ™ 4 hours with early stopping criterion on balanced accuracy. Inference on each task took fewer than an hour.

### 3.4 Ablation studies

We studied the effect of layer selection budget *K*, the number of gradient-ratio-selected layers on which LoRA adapters were applied. To isolate the effect of *K* from the choice of merging algorithm, all configurations used linear merging with *α*=1.0 throughout. We varied *K* ∈ {5, 7, 10, 15}, training all configurations with standard AdamW without projected gradient steps and with default LoRA initialization, so that the only factor under study was the layer selection budget. The baseline applied LoRA adapters uniformly across all transformer layers under the same optimizer and linear merging, serving as a configuration-matched upper bound on adapter coverage. Per-task balanced accuracy after merging and the corresponding fine-tune-to-merge gap are shown in Fig. 2.

**Figure 2:**
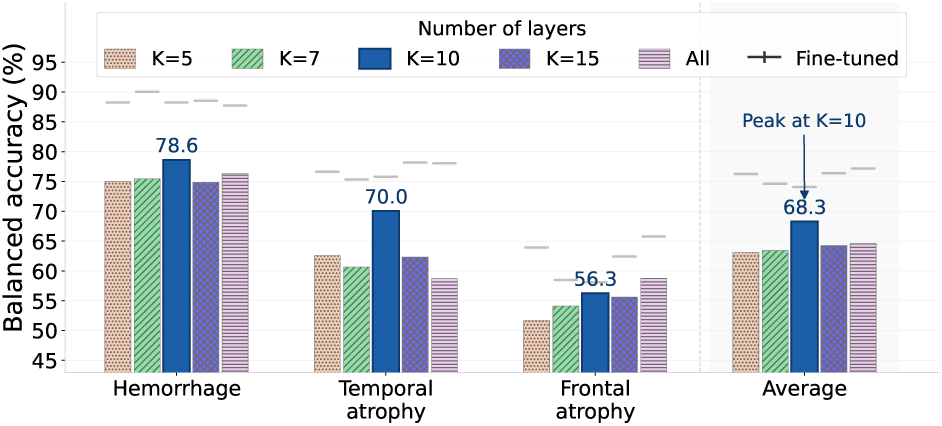
Layer selection ablation. Balanced accuracy of the linearly merged model as a function of the layer budget *K*. Per-task bars (Hemorrhage, Temporal atrophy, Frontal atrophy) are shown alongside the average. The fine-tuned performances per task are shown as dashed horizontal lines.

### 3.5 Performance metrics

Accuracy and F1-score were computed for each question format by evaluating correctness over the relevant answer options (e.g., { Yes, No} for Yes/No questions and {A, B, C} for MCQ). These metrics reflected the model’s question-answering proficiency given the specific prompt framing. Balanced accuracy was computed at the level of the underlying imaging evidence for each task; it quantified the model’s ability to distinguish MRI studies positive for the target imaging finding from normal studies, regardless of the question type. For comparison with baselines, we also reported the relative performance gap, defined as 100 * (Perf_merged_ ™ Perf_baseline_)*/*Perf_baseline_, where the baseline corresponds to either the jointly trained multi-task or individually fine-tuned model of the respective method.

## 4 Results

The zero-shot baseline confirmed the severity of the domain gap between natural image and multisequence brain MRI related question-answering tasks. Without adaptation, the model achieved near-chance balanced accuracy across all three imaging evidence tasks, and the degradation extended beyond class discrimination to question-answering tasks. This underscored that clinical imaging evidence cannot be recovered from pre-trained priors alone. Joint multi-task training provided a competitive ceiling for any merging strategy. We noted consistently weaker performance on temporal atrophy relative to the other tasks, an asymmetry that recurred across all methods and informed the per-task analysis below.

Averaged across both merging algorithms (Table 2(a)), GRAM yielded better answer-level accuracy and F1, exceeding both the all-layer baseline and Linear LoRA by up to 2.3% on BAcc, while adapting only *K*=5 transformer layers. Differences smaller than the across-seed standard deviation are reported as trends rather than significant gains. On question-level metrics, the average Acc and F1 of the GRAM-merged model exceeded the jointly trained multi-task ceiling by approximately 3.2% and 3.0% respectively; however, average BAcc, which is an imaging-evidence discrimination metric, remained about 8.8% below it, reflecting the residual cost of training-free merging on class-discrimination. Beyond raw performance, GRAM produced substantially more consistent merging outcomes across algorithms. With all-layer adapters, linear and TIES merging diverged by 14.4% on F1; under GRAM this divergence narrowed to 11.1%, achieving in training what TIES attempts post-hoc through its trim-and-elect mechanism. Under TIES merging individually, GRAM outperformed the all-layer configuration by more than 3% Acc and 2% F1 with fewer parameters. Under linear merging, GRAM matched the all-layer result within 1% Acc while improving BAcc by over 2%. This average advantage did not however hold uniformly for every algorithm-metric combination as on BAcc under TIES merging the all-layer baseline edged out GRAM suggesting that TIES’s post-hoc mechanism extracted a stronger signal from denser, less constrained adapters. The fine-tune-to-merge gap in BAcc (Table 2(a)), was 2.1% smaller under GRAM than the all-layer baseline and 3.3% smaller than Linear LoRA, confirming tighter coupling between the fine-tuned expert and its merged counterpart. The F1 gap further favored GRAM, recovering a net gain relative to the fine-tuned model compared to a net loss for Linear LoRA.

**Table 2:**
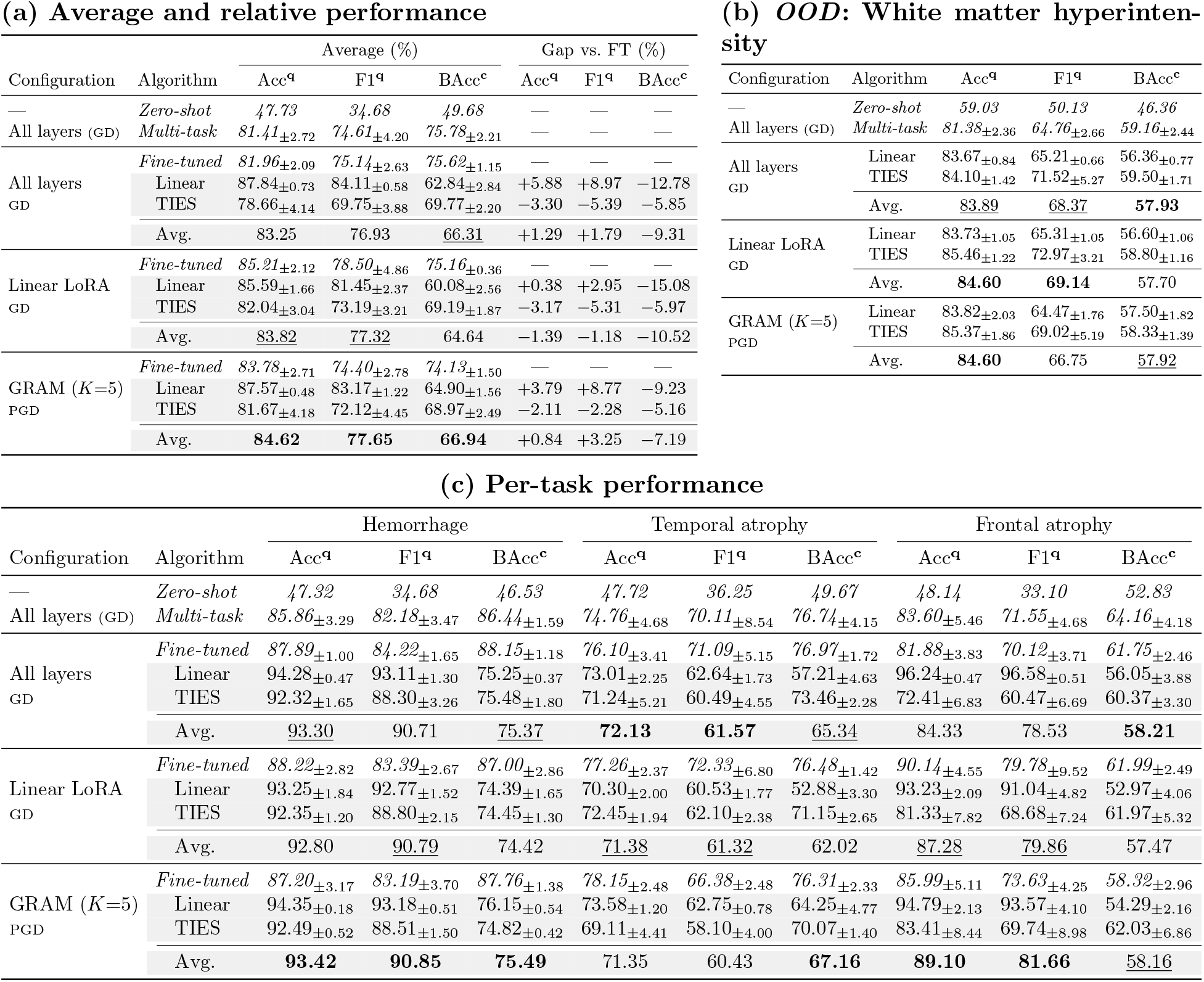
Performance on the NACC-VQA benchmark. Acc and F1 (marker **q**) are evaluated per question format; BAcc (marker **c**) is balanced accuracy at the imaging-evidence level. Results across 5 seeded runs are shown. In (a), Gap vs. FT is the merged model’s performance relative to the respective fine-tuned model. (b) Out-of-distribution (OOD) evaluation on white matter hyperintensity. (c) Performace across the three NACC-VQA tasks. Avg. denotes the mean over Linear and TIES merging algorithms for each configuration. GRAM uses *K*=5 gradient-ratio-selected layers with projected gradient descent.

The three tasks showed distinct interference patterns. GRAM led both baselines on all per-task average metrics for hemorrhage, with its merged Acc exceeding the jointly trained ceiling by over 7%, confirming that the visual signature of hemorrhage was robustly preserved under task composition. Temporal atrophy was the most challenging task across all methods, with average BAcc 9–15% below its multitask ceiling depending on configuration; GRAM reduced this gap relative to the all-layer baseline by nearly 2% on BAcc, consistent with the layer ablation (Section 3.4). Frontal atrophy exhibited constructive transfer under GRAM as the average Acc of merged models exceeded GRAM’s own individually fine-tuned expert by more than 3%, a pattern not observed in either baseline, attributable to shared feature extractors activated across the merged task vectors. A recurring observation across both frontal and temporal atrophy was higher variance in Table 2(c). This variance was not an artifact of merging as it was equally pronounced in the multi-task training condition, and reflected sensitivity of BAcc to the rare positive class under class imbalance. The OOD evaluation on white matter hyperintensity provided additional evidence of GRAM’s competitive generalization as the merged model exceeded the jointly trained ceiling by over 3% Acc while approaching it within 1.3% on BAcc.

Table 3 disaggregates results by question format. MCQ was the format where GRAM’s merged model most clearly surpassed multi-task training, suggesting that column-space projection particularly benefits the three-way discrimination required by this format. T/F and MPD showed GRAM meeting or exceeding the multi-task ceiling on hemorrhage, while Y/N and OSC were competitive on temporal atrophy. CV on hemorrhage remained the one consistent shortfall across both merging algorithms, indicating that sentence-level verification was partially disrupted when task vectors were composed.

**Table 3:** Performance per-question-type. Accuracy and F1 per question format. *Italic* rows are finetuned (FT) or reference models; indented rows are merged results (Linear,/ TIES) from the FT model above; Avg. is their mean. *All FT* : GD on all attention layers. *GRAM FT* : projected GD on top-*K* (*K*=5) gradient-ratio-selected layers. Numbers in row headers give test-sample counts per task (Hem.,Temp.,Front.).

|  | Method | Hem. |  | Temp. |  | Front. |  |
| --- | --- | --- | --- | --- | --- | --- | --- |
|  |  | Acc | F1 | Acc | F1 | Acc | F1 |
| (a) T/F (142/118/94) | *Zero-shot* | *35.92* | *28.03* | *51.69* | *44.30* | *57.45* | *51.87* |
|  | *Multi-task* | *84.23* | *83.97* | *74.75* | *74.56* | *83.19* | *83.04* |
|  | *All FT* | *86.48* | *86.40* | *72.37* | *72.29* | *81.06* | *80.99* |
|  | Linear | 96.20 | 96.19 | 73.05 | 72.93 | 94.89 | 94.89 |
|  | TIES | 84.65 | 84.20 | 64.58 | 64.29 | 68.30 | 64.99 |
|  | Avg. | 90.43 | 90.20 | 68.82 | 68.61 | 81.60 | 79.94 |
|  | *GRAM FT* | *87.32* | *87.08* | *75.08* | *74.98* | *85.32* | *85.30* |
|  | Linear | 97.18 | 97.18 | 73.39 | 73.29 | 94.04 | 94.03 |
|  | TIES | 87.75 | 87.55 | 65.42 | 64.60 | 77.45 | 75.40 |
|  | Avg. | **92.47** | **92.37** | **69.41** | **68.95** | **85.75** | **84.72** |
| (c) MPD (144/128/82) | *Zero-shot* | *52.08* | *37.80* | *51.56* | *36.71* | *50.00* | *33.33* |
|  | *Multi-task* | *82.92* | *82.87* | *72.97* | *72.39* | *72.44* | *72.25* |
|  | *All FT* | *81.94* | *81.93* | *70.31* | *70.03* | *68.78* | *68.52* |
|  | Linear | 96.25 | 96.25 | 66.41 | 66.38 | 97.56 | 97.56 |
|  | TIES | 93.89 | 93.88 | 62.03 | 61.46 | 37.56 | 36.40 |
|  | Avg. | 95.07 | 95.07 | 64.22 | 63.92 | 67.56 | 66.98 |
|  | *GRAM FT* | *78.61* | *78.61* | *73.44* | *73.29* | *79.02* | *78.57* |
|  | Linear | 95.14 | 95.14 | 67.34 | 66.98 | 97.32 | 97.32 |
|  | TIES | 95.56 | 95.56 | 62.03 | 61.75 | 78.29 | 78.20 |
|  | Avg. | **95.35** | **95.35** | **64.69** | **64.37** | **87.81** | **87.76** |
| (e) Y/N (217/193/139) | *Zero-shot* | *44.24* | *30.67* | *41.97* | *29.56* | *44.60* | *30.85* |
|  | *Multi-task* | *78.43* | *78.17* | *73.68* | *73.13* | *87.63* | *87.29* |
|  | *All FT* | *82.12* | *81.55* | *74.51* | *73.61* | *83.60* | *83.30* |
|  | Linear | 94.56 | 94.50 | 75.03 | 74.44 | 96.83 | 96.79 |
|  | TIES | 94.38 | 94.33 | 69.43 | 68.77 | 78.13 | 75.82 |
|  | Avg. | 94.47 | 94.42 | **72.23** | **71.61** | 87.48 | 86.31 |
|  | *GRAM FT* | *83.32* | *82.55* | *77.10* | *76.28* | *88.06* | *87.41* |
|  | Linear | 94.84 | 94.77 | 71.92 | 69.41 | 95.40 | 95.29 |
|  | TIES | 95.12 | 95.06 | 66.42 | 64.11 | 83.17 | 81.04 |
|  | Avg. | **94.98** | **94.92** | 69.17 | 66.76 | **89.29** | **88.17** |
| (b) OSC (130/120/84) | *Zero-shot* | *50.00* | *33.33* | *50.00* | *33.33* | *48.81* | *32.80* |
|  | *Multi-task* | *93.08* | *93.07* | *73.83* | *73.68* | *76.90* | *76.71* |
|  | *All FT* | *94.77* | *94.75* | *75.50* | *75.46* | *74.76* | *74.44* |
|  | Linear | 98.46 | 98.46 | 77.17 | 77.17 | 94.52 | 94.52 |
|  | TIES | 95.85 | 95.84 | 67.17 | 66.45 | 71.43 | 70.27 |
|  | Avg. | 97.16 | 97.15 | 72.17 | 71.81 | 82.98 | 82.40 |
|  | *GRAM FT* | *94.92* | *94.88* | *77.50* | *77.30* | *79.76* | *79.63* |
|  | Linear | 98.15 | 98.15 | 77.83 | 77.72 | 90.24 | 90.19 |
|  | TIES | 97.85 | 97.85 | 74.00 | 73.85 | 80.95 | 79.99 |
|  | Avg. | **98.00** | **98.00** | **75.92** | **75.79** | **85.60** | **85.09** |
| (d) CV (158/132/88) | *Zero-shot* | *51.90* | *38.38* | *48.48* | *35.07* | *50.00* | *33.33* |
|  | *Multi-task* | *97.59* | *97.59* | *76.97* | *76.94* | *96.36* | *96.32* |
|  | *All FT* | *99.87* | *99.87* | *82.12* | *81.97* | *97.95* | *97.95* |
|  | Linear | 87.34 | 87.34 | 71.21 | 71.21 | 96.14 | 96.14 |
|  | TIES | 89.62 | 89.62 | 72.88 | 72.22 | 90.45 | 90.10 |
|  | Avg. | **88.48** | **88.48** | 72.05 | 71.72 | 93.30 | 93.12 |
|  | *GRAM FT* | *97.59* | *97.57* | *84.55* | *84.53* | *97.73* | *97.73* |
|  | Linear | 87.72 | 87.72 | 73.64 | 73.44 | 95.68 | 95.68 |
|  | TIES | 87.72 | 87.72 | 75.91 | 75.89 | 96.82 | 96.82 |
|  | Avg. | 87.72 | 87.72 | **74.78** | **74.67** | **96.25** | **96.25** |
| (f) MCQ (48/53/24) | *Zero-shot* | *58.33* | *55.18* | *43.40* | *35.05* | *16.67* | *11.51* |
|  | *Multi-task* | *75.00* | *71.58* | *79.62* | *67.90* | *76.67* | *52.65* |
|  | *All FT* | *77.92* | *73.52* | *90.57* | *76.31* | *85.83* | *59.11* |
|  | Linear | 92.92 | 91.52 | 76.60 | 53.48 | 100.00 | 100.00 |
|  | TIES | 81.25 | 78.31 | 76.23 | 51.95 | 58.33 | 42.38 |
|  | Avg. | 87.09 | 84.92 | 76.42 | 52.72 | 79.17 | 71.19 |
|  | *GRAM FT* | *75.00* | *69.32* | *85.66* | *58.17* | *79.17* | *54.68* |
|  | Linear | 92.92 | 90.85 | 85.28 | 58.14 | 98.33 | 94.98 |
|  | TIES | 86.67 | 83.34 | 76.23 | 51.83 | 85.00 | 58.96 |
|  | Avg. | **89.80** | **87.10** | **80.76** | **54.99** | **91.67** | **76.97** |

Ablation on layer selection revealed that merged BAcc followed a non-monotonic pattern across *K*, peaking at *K*=10 with approximately 5% lift over *K*=5 and 4% over the all-layer baseline. Temporal atrophy drove the aggregate gain while hemorrhage remained stable across all values of *K*. Frontal atrophy achieved its best merged performance under the all-layer condition, indicating that its discriminative signal was more diffusely distributed across transformer layers. At *K*=10, GRAM achieved both the highest merged accuracy and the smallest fine-tune-to-merge gap (7.8%), compared with 17.3% at *K*=5 and 16.3% for the all-layer baseline. The performance gain at *K*=10 arose from reduced inter-task interference rather than from stronger individual adapters as the fine-tuned expert at *K*=10 achieved slightly lower individual BAcc (74.05%) than the all-layer baseline (77.17%), confirming that gradient-ratio-guided selection produced better-disentangled task vectors by restricting updates to task-discriminative layers. Notably, this optimal budget shifted to *K*=5 when projected gradient steps were applied.

## 5 Discussion

We present GRAM, a gradient-guided, subspace-constrained fine-tuning framework for parameter-efficient model merging in neuroimaging VLMs. Across the NACC-VQA benchmark, GRAM consistently improved the merged-model performance over standard all-layer LoRA, narrowing the fine-tune-to-merge gap and approaching the jointly trained multi-task ceiling without any joint re-training or cross-site data sharing. These results support our central hypothesis that the placement and geometry of task-specific updates govern the success of model composition in clinically meaningful imaging settings.

A distinctive contribution of GRAM is gradient ratio, a class-contrastive per-layer importance metric that contrasts class-level gradient magnitudes to identify layers. Layer importance for adapter finetuning has received limited systematic attention, where existing approaches largely apply LoRA uniformly across all transformer layers without accounting for the heterogeneous sensitivity of different layers to domain-specific clinical findings. The gradient ratio addressed this gap directly, and the ablation study identified a compact, task-discriminative sub-space that cannot be recovered by uniform adapter placement. Complementing the gradient ratio, the column-space projected gradient descent constrains task-specific LoRA updates to respect the geometric structure already encoded in the pre-trained weights, encouraging task vectors to remain aligned. This structured optimization produces adapters that are inherently more amenable to post-hoc composition. To our knowledge, GRAM also represents the first systematic application of model merging to VLMs for neuroimaging, a domain where multi-sequence volumetric MRI and class-imbalance pose different challenges than the natural-image benchmarks on which merging methods are typically developed and evaluated. Our proposed framework has limitations. The SVD decomposition required for column-space projected gradient descent incurs non-trivial computational overhead per selected layer. Although the decomposition is computed once and cached for reuse, this upfront cost can become a practical bottleneck with larger base models. Additionally, the present study evaluates GRAM exclusively on the NACC cohort, a single retrospective dataset collected under relatively uniform acquisition protocols, and the gradient ratio assumes labeled positive and normal subsets per task to compute the class contrast.

Collectively, these findings position GRAM as a step towards a collaborative neuroimaging model. Independent clinical institutions may fine-tune VLMs on their respective patient populations for distinct clinical indications, each operating exclusively on locally held imaging data. By contributing only the resulting adapter weights rather than patient records, these independently trained specialists can be composed into a unified model that integrates complementary clinical knowledge. GRAM’s gradient-ratio-guided layer selection and subspace-constrained fine-tuning makes this composition reliable by reducing destructive interference during merging. This directly addresses one barrier to large-scale neuroimaging models, where the difficulty of aggregating data across institutions subject to differing regulatory frameworks and data governance policies. Extending this framework to a broader federation of sites and disease domains can be a principled path toward a large-scale multi-task neuroimaging model.

## Data Availability

Data was obtained from https://www.naccdata.org/.

## Footnotes

1 Under LoRA-based adaptation, since *θ*_t_ = *θ*_0_ + Δ*θ*_t_, the task vector becomes *τ*_t_ = Δ*θ*_t_.

## Notes

### Competing Interest Statement

V.B.K. is a co-founder of Cognimark, Inc. He also serves on the scientific advisory board of Altoida Inc. The remaining authors declare no competing interests.

### Author Declarations

The study used ONLY openly available human data that were originally located at: https://www.naccdata.org/.

